# Effectiveness of bivalent mRNA booster vaccination against SARS-CoV-2 Omicron infection in the Netherlands, September to December 2022

**DOI:** 10.1101/2023.02.08.23285643

**Authors:** Anne J. Huiberts, Brechje de Gier, Christina E. Hoeve, Hester E. de Melker, Susan J.M. Hahné, Gerco den Hartog, Janneke H.H.M. van de Wijgert, Susan van den Hof, Mirjam J. Knol

## Abstract

We used data of 32,542 prospective cohort study participants who previously received primary and one or two monovalent booster COVID-19 vaccinations. Between 26 September and 19 December 2022, relative effectiveness of bivalent Original/Omicron BA.1 vaccination against self-reported Omicron SARS-CoV-2 infection was 31% in 18-59-year-olds and 14% in 60-85-year-olds. Protection was higher after prior Omicron infection than after bivalent vaccination without prior infection. Although bivalent booster vaccination increases protection against COVID-19 hospitalizations, we found limited added benefit in preventing SARS-CoV-2 infection.

---

The SARS-CoV-2 Omicron variant has been dominant in Europe since January 2022, causing large waves of infections because of high transmissibility and escape from vaccine- and infection-induced immunity [1]. Bivalent mRNA vaccines targeting the Omicron BA.1 subvariant and the original strain of SARS-CoV-2 were available as booster vaccination for all individuals aged 12 years and older in the Netherlands since September 19, 2022. Individuals aged ≥ 60 years, medical risk groups and health care workers were actively invited. We present estimates of the relative effectiveness of bivalent Omicron BA.1-targeted vaccination against self-reported SARS-CoV-2 Omicron infection between 26 September and 19 December 2022 among adults who had previously received primary vaccination and one or two monovalent booster vaccinations.

## Study population

We used data from 32,542 participants of an ongoing prospective cohort study (VASCO) among community-dwelling Dutch adults aged 18-85 years who are followed with questionnaires and 6-monthly serum samples [2, 3]. Follow-up started on 26 September 2022 (one week after the start of the bivalent booster vaccination program), or three months after the last monovalent vaccination or last prior infection (occurring before 26 September 2022), whichever came last. This is in line with vaccination policy, where individuals are eligible for a bivalent vaccine three months after vaccination or infection. Follow-up ended on 19 December 2022, at the date of first positive SARS-CoV-2 test or at the date of last completed follow-up questionnaire, whichever came first. Participants aged 18-59 years who previously received a primary vaccination series and one monovalent booster vaccination were included in the analysis (n=12,988). Participants aged 60-85 years who previously received a primary vaccination series and one (n=8,963) or two (n=10,591) monovalent booster vaccinations were included in the analysis (n=19,554). In total, 5,504 (42.4%) 18-59-year-olds and 11,900 (60.9%) 60-85-year-olds received a bivalent vaccine (**Table 1**). Prior SARS-CoV-2 infection, based on self-report or presence of SARS-CoV-2 Nucleoprotein-specific antibodies [2], was present in 9,605 (73.4%) of 18-59-year-olds and 10,898 (55.7%) of 60-85-year-olds (**Table 1**). Participants who received the bivalent booster vaccine were older (median age 51 vs. 48 in 18-59-year-olds) and more often had a medical risk condition (26.5% vs 18.0% in 18-59-year-olds; 41.9% vs 38.2% in 60-85-year-olds) than participants who did not receive a booster during the study period. Among 60-85-year-olds, the bivalent booster vaccine recipients more frequently received two prior monovalent booster vaccinations than the non-recipients (58.2% vs 47.9%).

**Table 1.**
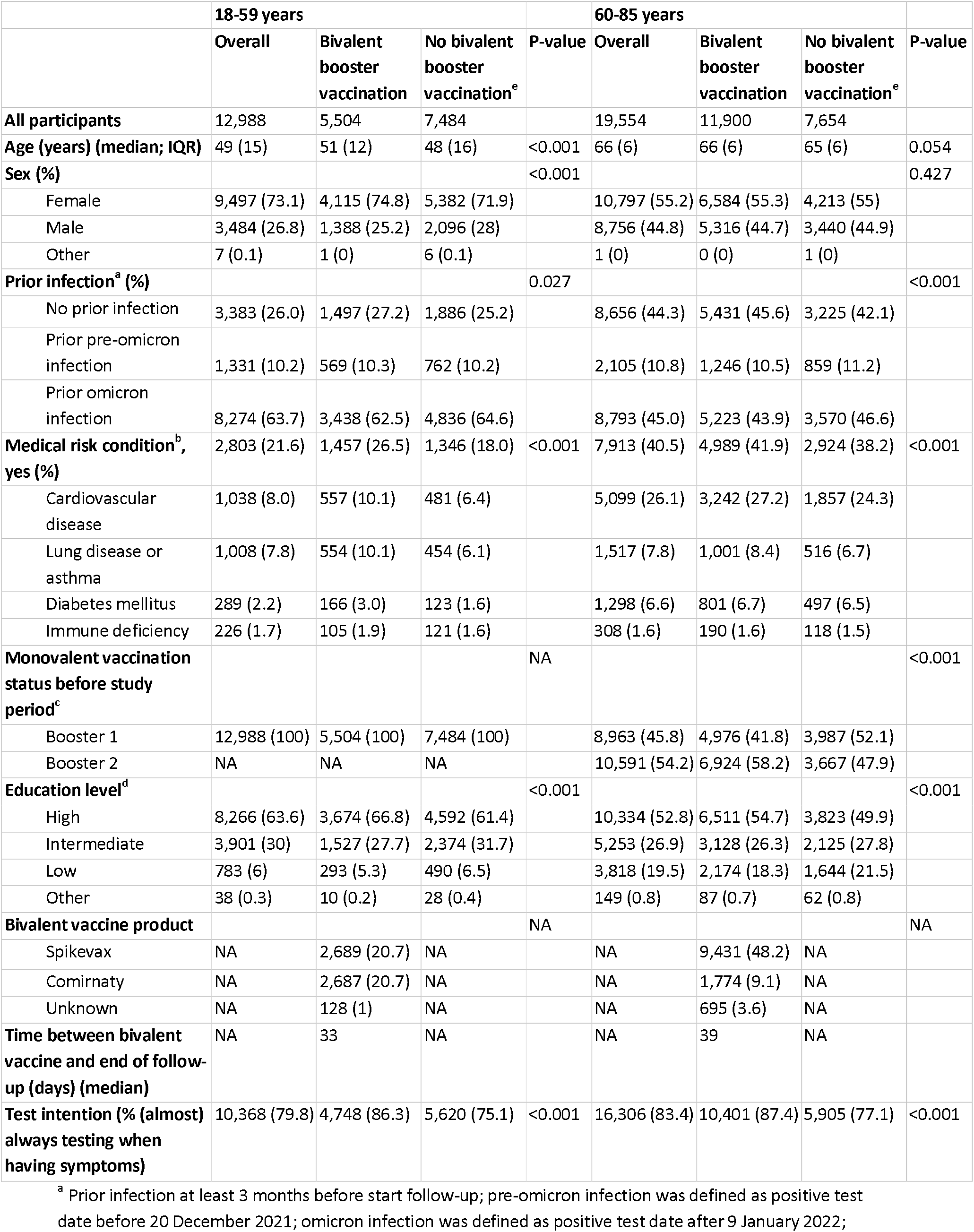
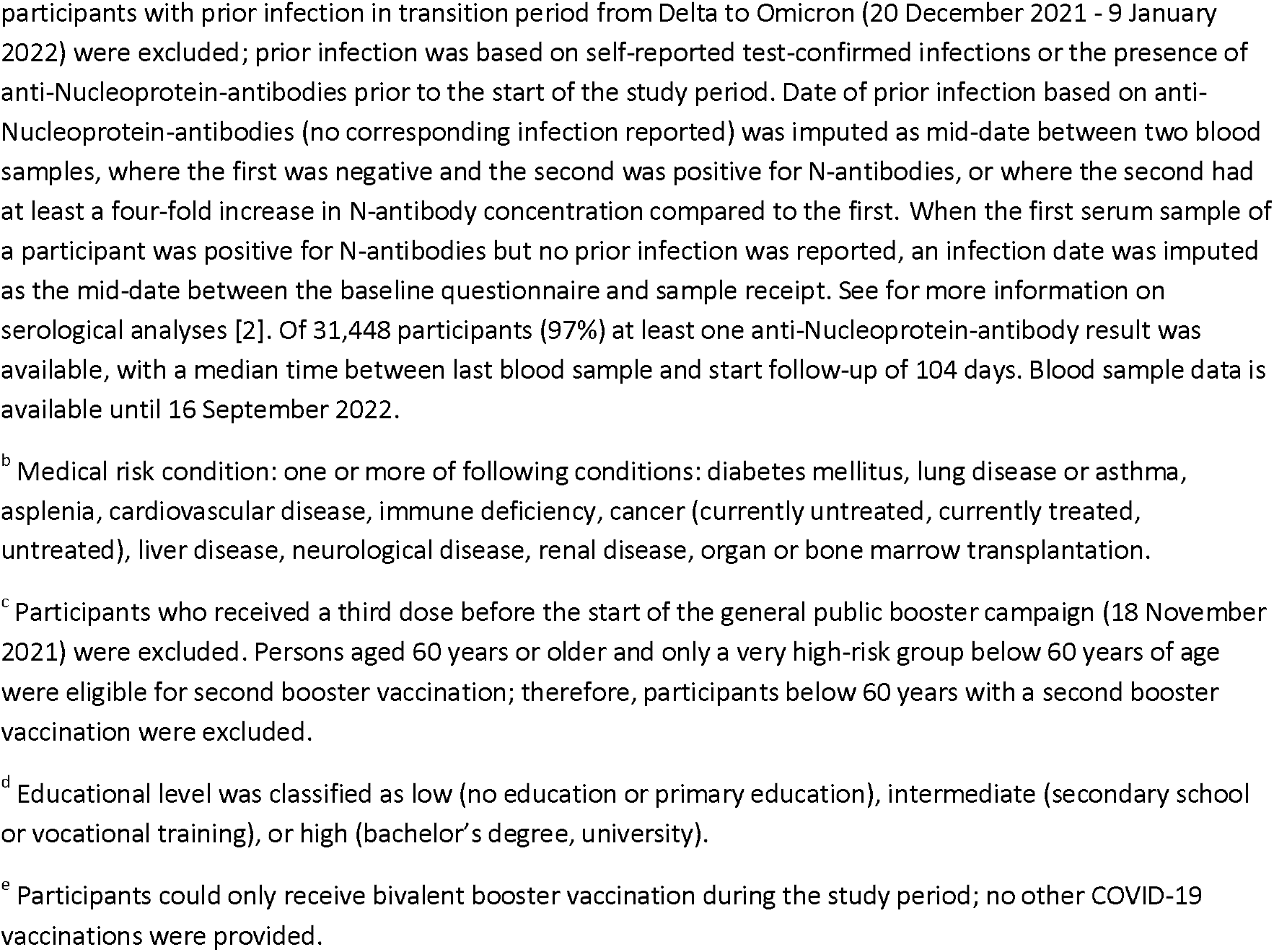
Characteristics of participants included in the analysis

|  | 18-59 years |  |  |  | 60-85 years |  |  |  |
| --- | --- | --- | --- | --- | --- | --- | --- | --- |
|  | Overall | Bivalent booster vaccination | No bivalent booster vaccination<sup>e</sup> | P-value | Overall | Bivalent booster vaccination | No bivalent booster vaccination<sup>e</sup> | P-value |
| **All participants** | 12,988 | 5,504 | 7,484 |  | 19,554 | 11,900 | 7,654 |  |
| **Age (years) (median; IQR)** | 49 (15) | 51 (12) | 48 (16) | <0.001 | 66 (6) | 66 (6) | 65 (6) | 0.054 |
| **Sex (%)** |  |  |  | <0.001 |  |  |  | 0.427 |
| Female | 9,497 (73.1) | 4,115 (74.8) | 5,382 (71.9) |  | 10,797 (55.2) | 6,584 (55.3) | 4,213 (55) |  |
| Male | 3,484 (26.8) | 1,388 (25.2) | 2,096 (28) |  | 8,756 (44.8) | 5,316 (44.7) | 3,440 (44.9) |  |
| Other | 7 (0.1) | 1 (0) | 6 (0.1) |  | 1 (0) | 0 (0) | 1 (0) |  |
| **Prior infection<sup>a</sup> (%)** |  |  |  | 0.027 |  |  |  | <0.001 |
| No prior infection | 3,383 (26.0) | 1,497 (27.2) | 1,886 (25.2) |  | 8,656 (44.3) | 5,431 (45.6) | 3,225 (42.1) |  |
| Prior pre-omicron infection | 1,331 (10.2) | 569 (10.3) | 762 (10.2) |  | 2,105 (10.8) | 1,246 (10.5) | 859 (11.2) |  |
| Prior omicron infection | 8,274 (63.7) | 3,438 (62.5) | 4,836 (64.6) |  | 8,793 (45.0) | 5,223 (43.9) | 3,570 (46.6) |  |
| **Medical risk condition<sup>b</sup>, yes (%)** | 2,803 (21.6) | 1,457 (26.5) | 1,346 (18.0) | <0.001 | 7,913 (40.5) | 4,989 (41.9) | 2,924 (38.2) | <0.001 |
| Cardiovascular disease | 1,038 (8.0) | 557 (10.1) | 481 (6.4) |  | 5,099 (26.1) | 3,242 (27.2) | 1,857 (24.3) |  |
| Lung disease or asthma | 1,008 (7.8) | 554 (10.1) | 454 (6.1) |  | 1,517 (7.8) | 1,001 (8.4) | 516 (6.7) |  |
| Diabetes mellitus | 289 (2.2) | 166 (3.0) | 123 (1.6) |  | 1,298 (6.6) | 801 (6.7) | 497 (6.5) |  |
| Immune deficiency | 226 (1.7) | 105 (1.9) | 121 (1.6) |  | 308 (1.6) | 190 (1.6) | 118 (1.5) |  |
| **Monovalent vaccination status before study period<sup>c</sup>** |  |  |  | NA |  |  |  | <0.001 |
| Booster 1 | 12,988 (100) | 5,504 (100) | 7,484 (100) |  | 8,963 (45.8) | 4,976 (41.8) | 3,987 (52.1) |  |
| Booster 2 | NA | NA | NA |  | 10,591 (54.2) | 6,924 (58.2) | 3,667 (47.9) |  |
| **Education level<sup>d</sup>** |  |  |  | <0.001 |  |  |  | <0.001 |
| High | 8,266 (63.6) | 3,674 (66.8) | 4,592 (61.4) |  | 10,334 (52.8) | 6,511 (54.7) | 3,823 (49.9) |  |
| Intermediate | 3,901 (30) | 1,527 (27.7) | 2,374 (31.7) |  | 5,253 (26.9) | 3,128 (26.3) | 2,125 (27.8) |  |
| Low | 783 (6) | 293 (5.3) | 490 (6.5) |  | 3,818 (19.5) | 2,174 (18.3) | 1,644 (21.5) |  |
| Other | 38 (0.3) | 10 (0.2) | 28 (0.4) |  | 149 (0.8) | 87 (0.7) | 62 (0.8) |  |
| **Bivalent vaccine product** |  |  |  | NA |  |  |  | NA |
| Spikevax | NA | 2,689 (20.7) | NA |  | NA | 9,431 (48.2) | NA |  |
| Comirnaty | NA | 2,687 (20.7) | NA |  | NA | 1,774 (9.1) | NA |  |
| Unknown | NA | 128 (1) | NA |  | NA | 695 (3.6) | NA |  |
| **Time between bivalent vaccine and end of follow-up (days) (median)** | NA | 33 | NA |  | NA | 39 | NA |  |
| **Test intention (% (almost always testing when having symptoms))** | 10,368 (79.8) | 4,748 (86.3) | 5,620 (75.1) | <0.001 | 16,306 (83.4) | 10,401 (87.4) | 5,905 (77.1) | <0.001 |

<sup>a</sup> Prior infection at least 3 months before start follow-up; pre-omicron infection was defined as positive test date before 20 December 2021; omicron infection was defined as positive test date after 9 January 2022;
participants with prior infection in transition period from Delta to Omicron (20 December 2021 - 9 January 2022) were excluded; prior infection was based on self-reported test-confirmed infections or the presence of anti-Nucleoprotein-antibodies prior to the start of the study period. Date of prior infection based on anti-Nucleoprotein-antibodies (no corresponding infection reported) was imputed as mid-date between two blood samples, where the first was negative and the second was positive for N-antibodies, or where the second had at least a four-fold increase in N-antibody concentration compared to the first. When the first serum sample of a participant was positive for N-antibodies but no prior infection was reported, an infection date was imputed as the mid-date between the baseline questionnaire and sample receipt. See for more information on serological analyses [2]. Of 31,448 participants (97%) at least one anti-Nucleoprotein-antibody result was available, with a median time between last blood sample and start follow-up of 104 days. Blood sample data is available until 16 September 2022.
<sup>b</sup> Medical risk condition: one or more of following conditions: diabetes mellitus, lung disease or asthma, asplenia, cardiovascular disease, immune deficiency, cancer (currently untreated, currently treated, untreated), liver disease, neurological disease, renal disease, organ or bone marrow transplantation.
<sup>c</sup> Participants who received a third dose before the start of the general public booster campaign (18 November 2021) were excluded. Persons aged 60 years or older and only a very high-risk group below 60 years of age were eligible for second booster vaccination; therefore, participants below 60 years with a second booster vaccination were excluded.
<sup>d</sup> Educational level was classified as low (no education or primary education), intermediate (secondary school or vocational training), or high (bachelor's degree, university).
<sup>e</sup> Participants could only receive bivalent booster vaccination during the study period; no other COVID-19 vaccinations were provided.

## Incidence of SARS-CoV-2 infection

During the study period, 3,005 SARS-CoV-2 infections, based on a positive SARS-CoV-2 PCR or (self-administered) antigen test, were reported by the participants. The reported incidence in September and October 2022 was high (**Figure 1**), consistent with national data from syndromic and wastewater surveillance [4, 5]. The incidence was highest among participants without any prior infection, lower among participants with a prior pre-Omicron infection, and lowest among participants with a prior Omicron infection. During most of the study period, the incidence was lower among participants who did than among those who did not receive a bivalent booster vaccine. However, it is important to note that the number of participants with a bivalent vaccine was low in the beginning of the study period (**Figure S1**) and thus the incidence in these participants was based on a low number of infections.

**Figure 1.**
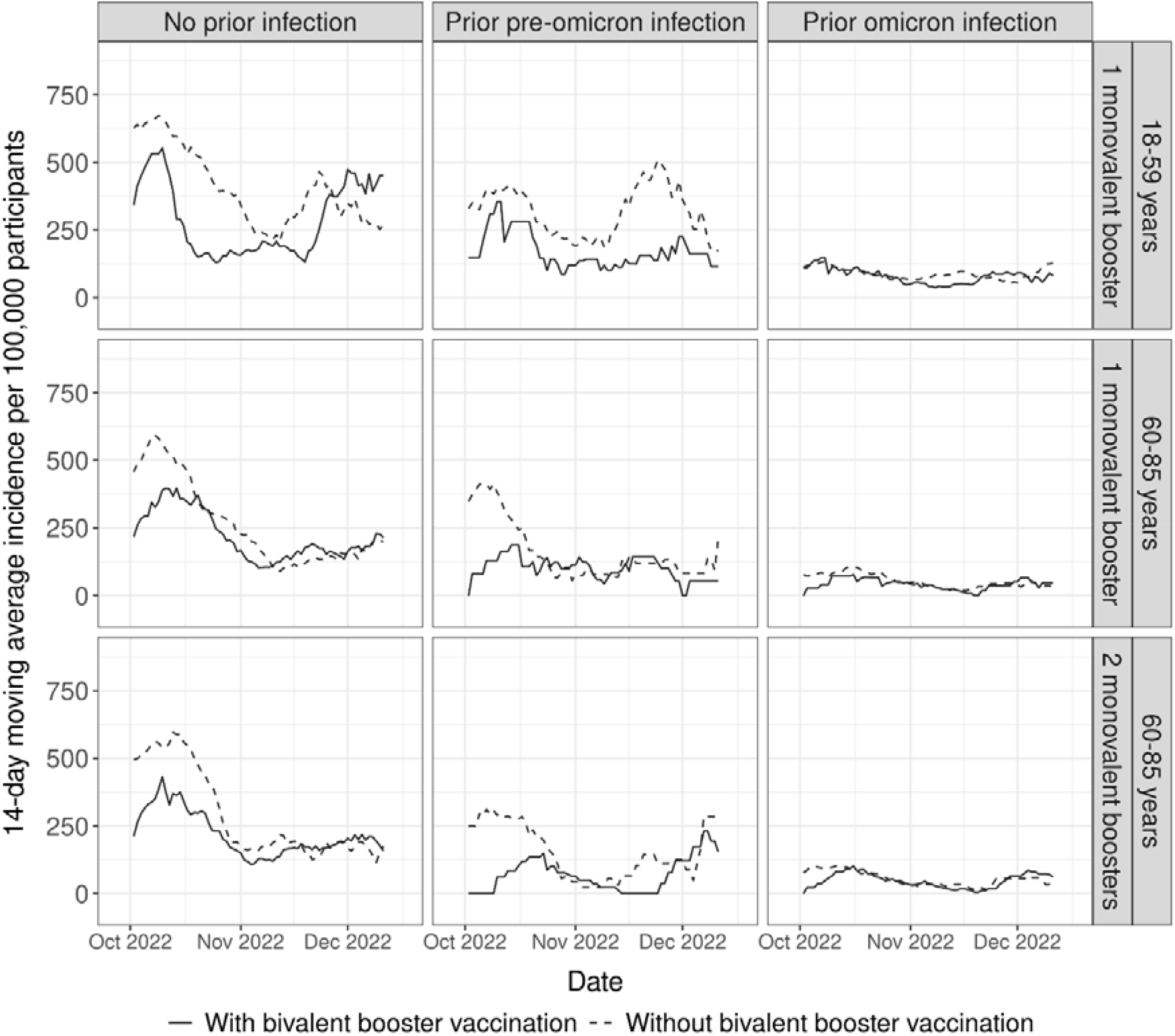
14-day moving average of number of infections reported per 100,000 participants by age group, prior infection status and vaccination status from 26 September to 19 December 2022

## Relative vaccine effectiveness

To estimate effectiveness of bivalent vaccination relative to receiving the primary vaccination series and one or two monovalent booster vaccinations, we used Cox proportional hazard models with calendar time as underlying time scale and bivalent vaccination as time-varying exposure. Estimates were adjusted for age group, sex, education level and presence of a medical risk condition. We present stratified estimates by infection history and an overall estimate additionally adjusted for infection history. The 7 person-days after bivalent vaccine administration were excluded. All analyses were done using R version 4.2.2 and packages Epi and survival.

Among 18-59-year-olds who received primary vaccination and one monovalent booster, the overall relative effectiveness of bivalent vaccination against infection was 31% (95%CI: 18-42). Among participants with prior Omicron infection the relative effectiveness of a bivalent booster was lower (20%; 95%CI: -7-40) (**Figure 2, Table S1**). Among 60-85-year-olds who received primary vaccination and one or two monovalent booster vaccinations, relative effectiveness was 14% (95%CI: 3-24) and 6% (95%CI: -30-31) among participants with prior Omicron infection.

**Figure 2.**
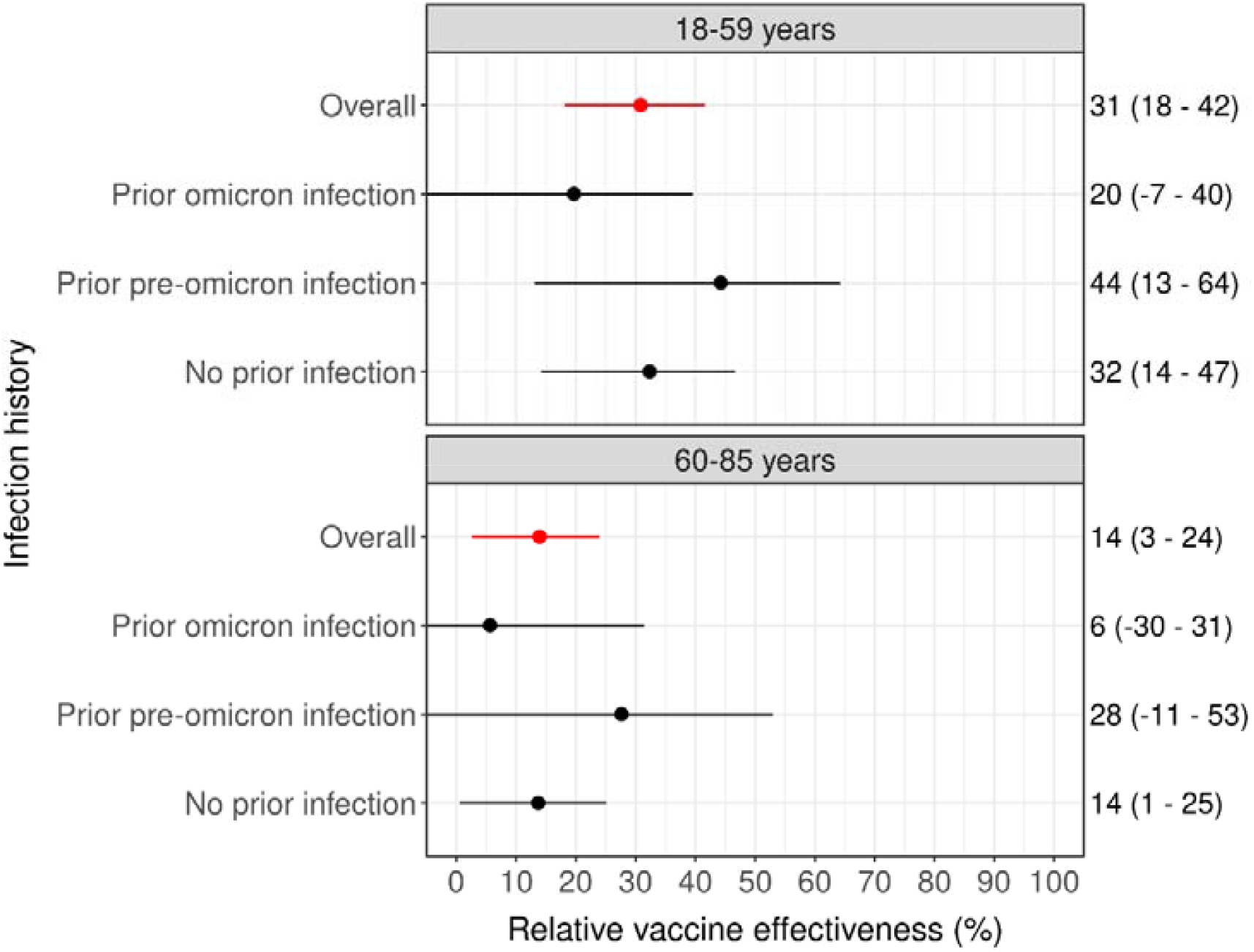
Relative vaccine effectiveness and 95% confidence interval of bivalent vaccine overall and stratified by infection history and by age group from 26 September 2022 to 19 December 2022. ^a^ Adjusted for age group (18-39, 40-59, 60-69, 70-85), sex, education level and presence of a medical risk condition; overall estimates were additionally adjusted for infection history.

Estimates among 60-85-year-olds were similar to the main estimate across different stratified analyses and sensitivity analyses (**Figure 3**). Among 18-59-year-olds, stratification by bivalent vaccine product showed higher relative effectiveness of Spikevax (Moderna) than Comirnaty (BioNtech/Pfizer) bivalent vaccine; of note, Spikevax was only given to individuals aged 45 years and older and therefore the median age in Spikevax-recipients was higher than in Comirnaty-recipients (54 vs. 43) (**Figure 3**).

**Figure 3.**
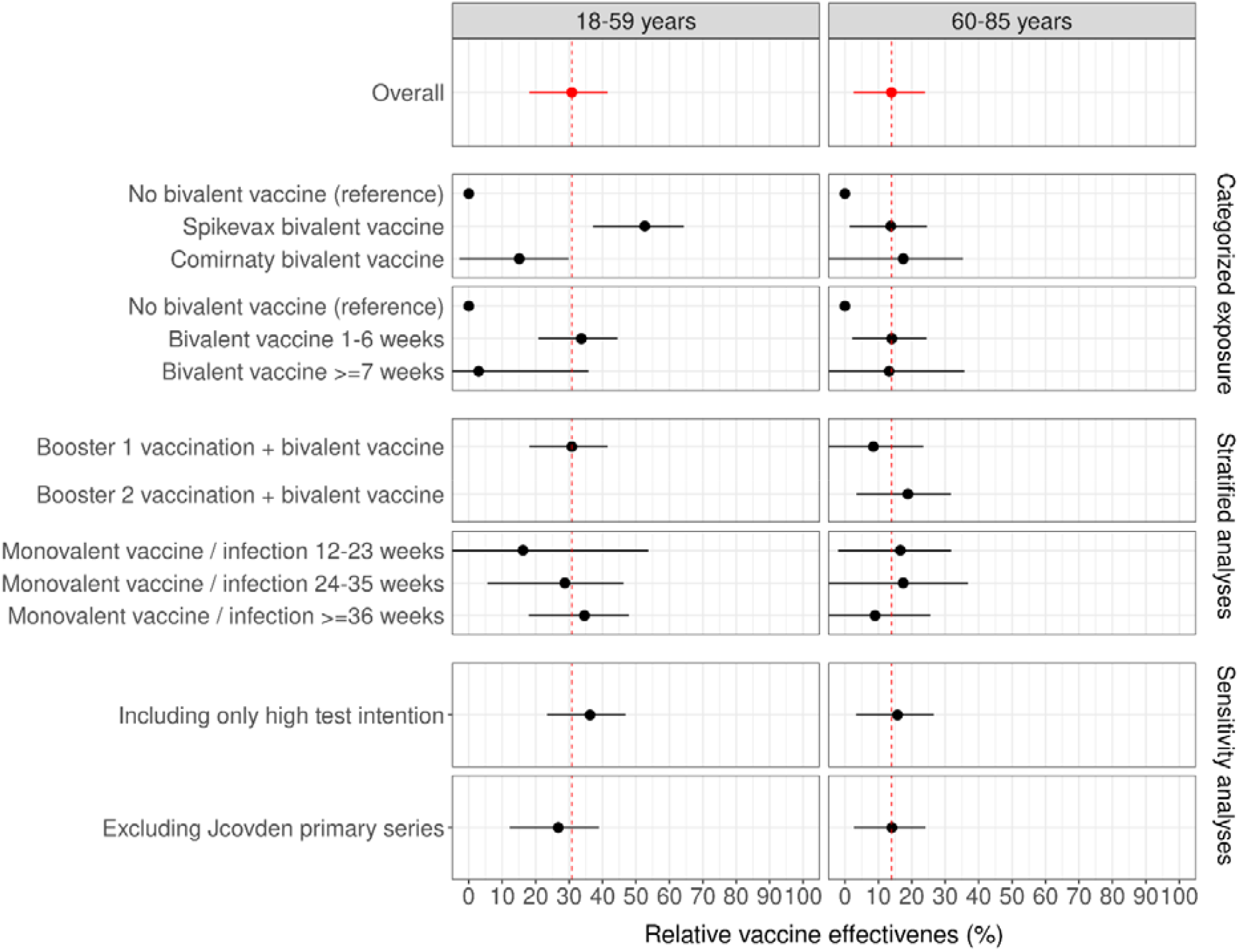
Stratified and sensitivity analyses by age group from 26 September 2022 to 19 December 2022 ^a^ Adjusted for age group (18-39, 40-59, 60-69, 70-85), sex, education level, presence of a medical risk condition, and infection history. Categorized exposure = exposure categorized by 1) vaccine product and 2) time since bivalent vaccination; Stratified analyses = analyses stratified by 1) number of monovalent booster doses before start of follow-up and 2) time since last monovalent booster at start follow-up; Sensitivity analyses = analyses performed in 1) only persons with a high intention to test during follow-up and 2) excluding all individual with a Jcovden primary series.

In participants aged 18-59 years, compared to those without bivalent vaccination and without prior infection, relative effectiveness of bivalent vaccination among participants without prior infection (37%; 95%CI: 21-50) was similar to relative protection from a prior pre-Omicron infection and no bivalent vaccination (34%; 95%CI: 21-44), while relative protection from a prior Omicron infection with or without bivalent vaccination was substantially higher (80-83%) (**Table S2**). Similarly, participants aged 60-85 years showed higher relative protection from prior Omicron infection with or without bivalent vaccination (82%) than from bivalent vaccination (14%; 95%CI 1-25) or prior pre-Omicron infection (43%; 95%CI: 32-52).

## Discussion and conclusion

We found that Original/Omicron BA.1-targeted bivalent vaccination gave an overall relative vaccine effectiveness against Omicron SARS-CoV-2 infection of 31% in 18-59-year-olds and 14% in 60-85-year-olds who had previously received the primary vaccination series and at least one booster vaccination, adjusted for infection history.

Our data showed higher protection of prior Omicron infection compared to the protection of bivalent vaccination among persons without prior infection, even though time since prior Omicron infection was longer than time since bivalent vaccination. This is consistent with a recent preprint estimating higher and longer protection after a breakthrough infection compared to booster vaccination [6]. In general, a combination of vaccination and infection, i.e. hybrid immunity, has been shown to provide better protection against infection than vaccination alone [7, 8]. We did, however, find a similar effect of prior pre-Omicron infection and bivalent vaccination without prior infection. Probably because time since pre-Omicron infection was significantly longer than time since bivalent vaccination.

Estimates of (relative) effectiveness of bivalent vaccination against infection are scarce. A recent study from the US reported slightly higher estimates against infection by the BA.4/BA.5-targeted bivalent vaccine (46%, 38%, and 36% 6-7 months after last monovalent dose in individuals aged 18-49 years, 50-64 and ≥65 years) [9]. However, these estimates were not stratified by or adjusted for infection history. A preprint from the Nordic countries reported a relative effectiveness against hospitalization of 75% for the BA.1-targeted bivalent vaccine in individuals aged ≥50 years [10]. Dutch surveillance data reported a relative risk reduction of 45% in 40-59-year-olds and 58% of BA.1-targeted bivalent vaccination in individuals aged ≥60 years with at least one prior monovalent vaccination [11].

The VASCO cohort participants were given SARS-CoV-2 self-administered antigen tests free of charge, and we were not dependent on the SARS-CoV-2 testing infrastructure. Additionally, serological data enabled us to detect prior untested (asymptomatic) infections. Estimates can be confounded through differences in (time-varying) factors between participants who did and did not receive a bivalent booster vaccine, including test frequency and differences in exposure through behaviour. Participants who received a bivalent booster vaccine had a slightly higher intention to test, but restricting to participants with high test intention did not change our estimates. Since we investigated participants who already received monovalent booster vaccination only and COVID-19 measures were limited during the study period, differences in SARS-CoV-2 exposure between bivalent vaccine recipients and non-recipients will likely be limited.

The bivalent booster vaccination campaign has shown benefit in reducing COVID-19 hospitalizations, which is especially important for those at increased risk including elderly and those with a medical risk condition. However, we found limited added protection of bivalent vaccination in preventing SARS-CoV-2 Omicron infection among persons who received primary vaccination and one or two monovalent booster vaccinations. Especially in persons with prior Omicron infection, the added benefit seems low.

## Data Availability

All data produced in the present study are available in aggregated and anonymized form upon reasonable request to the authors.

