## Supplementary material for "Effectiveness of bivalent mRNA booster vaccination against SARS-CoV-2 Omicron infection in the Netherlands, September to December 2022": Table S

**Figure S1**. Number of participants and reported SARS-CoV-2 infections per vaccination status over time stratified by infection history and age group from 26 September 2022 to 19 December 2022

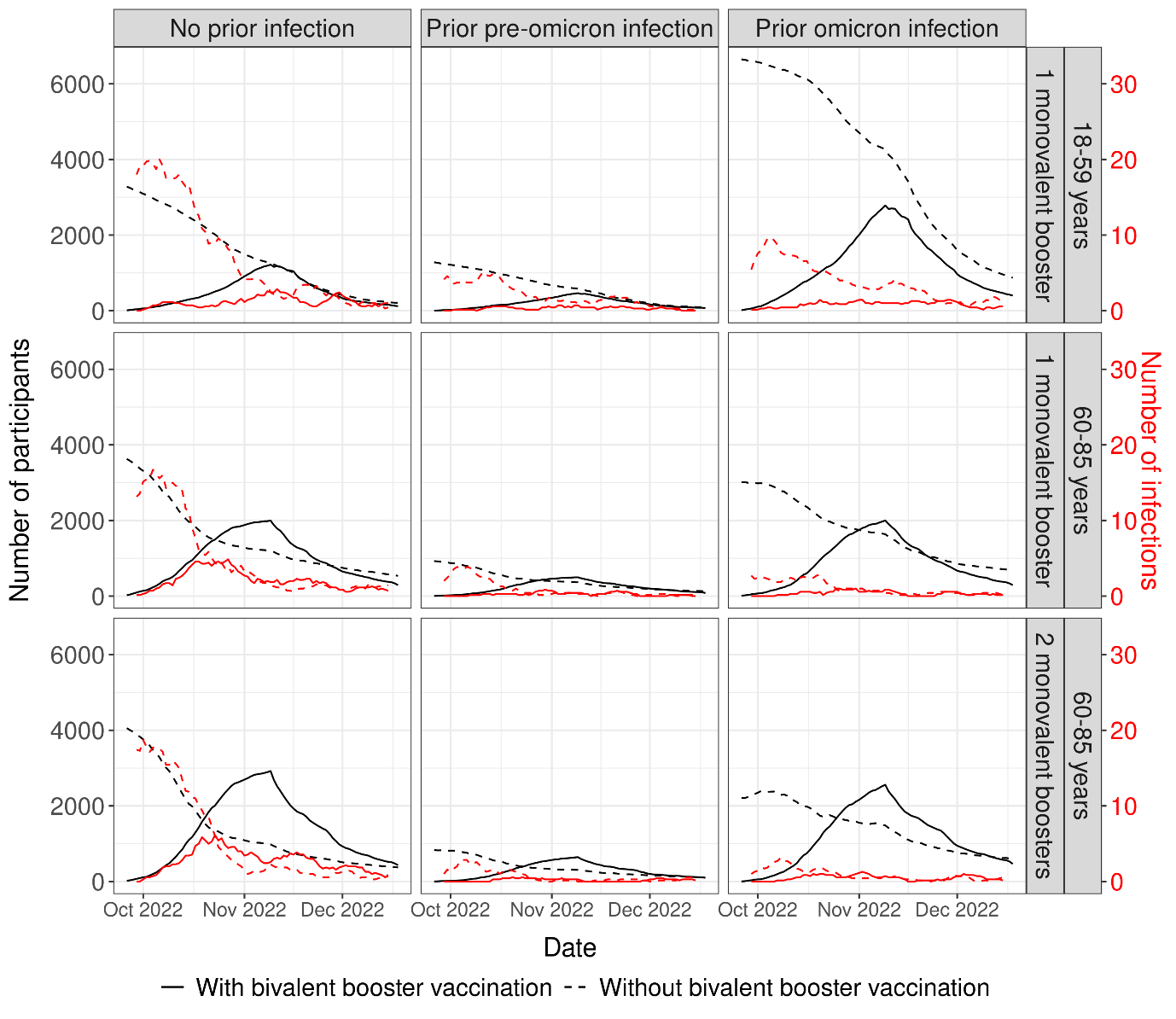

**Table S1**. Incidence rate and relative vaccine effectiveness of bivalent vaccine stratified by infection history and age group from 26 September 2022 to 19 December 2022

|  | **Person-weeks** | **Number of infections** | **Rate (per 1,000 weeks)** | **Adjusted^a^ relative vaccine effectiveness (95% CI)** |
| --- | --- | --- | --- | --- |
| **18-59 years** |  |  |  |  |
| Overall |  |  |  | 31 (18-42) |
| No prior infection | 5,882 | 98 | 16.7 | 32 (14-47) |
| Prior pre-omicron infection | 2,419 | 26 | 10.7 | 44 (13-64) |
| Prior omicron infection | 14,242 | 66 | 4.6 | 20 (-7-40) |
| **60-85 years** |  |  |  |  |
| Overall |  |  |  | 14 (3-24) |
| No prior infection | 28,045 | 381 | 13.6 | 14 (1-25) |
| Prior pre-omicron infection | 6,295 | 35 | 5.6 | 28 (-11-53) |
| Prior omicron infection | 24,458 | 68 | 2.8 | 6 (-30-31) |

^a^ Adjusted for age group (18-39, 40-59, 60-69, 70-85), sex, education level and chronic condition. Overall estimates were additionally adjusted for infection history.

**Table S2**. Effectiveness^a^ of bivalent vaccination and prior infection status stratified by age group from 26 September 2022 to 19 December 2022

|  | **No prior infection** | **Prior pre-omicron infection** | **Prior omicron infection** |
| --- | --- | --- | --- |
| **18-59 years** |  |  |  |
| No bivalent vaccination | Reference | 34 (21-44) | 80 (77-83) |
| Bivalent vaccination | 37 (21-50) | 60 (41-73) | 83 (78-87) |
| **60-85 years** |  |  |  |
| No bivalent vaccination | Reference | 43 (32-52) | 82 (79-85) |
| Bivalent vaccination | 14 (1-25) | 63 (48-74) | 82 (76-86) |

^a^ Adjusted for age group (18-39, 40-59, 60-69, 70-85), sex, education level and chronic condition

**Table S3**. Incidence rate and relative vaccine effectiveness of bivalent vaccine in participants who (almost) always test in case of COVID-19 symptoms stratified by infection history and age group from 26 September 2022 to 19 December 2022

|  | **Person-weeks** | **Number of infections** | **Rate (per 1,000 weeks)** | **Adjusted^a^ relative vaccine effectiveness (95% CI)** |
| --- | --- | --- | --- | --- |
| **18-59 years** |  |  |  |  |
| Overall |  |  |  | 36 (23-47) |
| No prior infection | 5,029 | 83 | 16.5 | 40 (22-53) |
| Prior pre-omicron infection | 2,004 | 22 | 11.0 | 47 (14-67) |
| Prior omicron infection | 11,956 | 58 | 4.9 | 23 (-5-44) |
| **60-85 years** |  |  |  |  |
| Overall |  |  |  | 16 (3-27) |
| No prior infection | 23,615 | 330 | 14.0 | 15 (1-28) |
| Prior pre-omicron infection | 5,106 | 32 | 6.3 | 25 (-19-53) |
| Prior omicron infection | 20,667 | 56 | 2.7 | 11 (-26-38) |

^a^ Adjusted for age group (18-39, 40-59, 60-69, 70-85), sex, education level and chronic condition. Overall estimates were additionally adjusted for infection history.
